# Artificial Intelligence Neural Network Consistently Interprets Lung Ultrasound Artifacts in Hospitalized Patients: A Prospective Observational Study

**DOI:** 10.1101/2023.03.02.23286687

**Authors:** Thomas H. Fox, Gautam R. Gare, Laura E. Hutchins, Victor S. Perez, Ricardo Rodriguez, David L. Smith, Francisco X. Brito-Encarnacion, Raman Danrad, Hai V. Tran, Peter B. Lowery, David J. Montgomery, Kevin A. Zamorra, Amita Krishnan, John M. Galeotti, Bennett P. deBoisblanc

**Author notes:** Please direct all correspondence to Dr. Bennett P. deBoisblanc by email at.

## Abstract

**Background:** Interpretation of lung ultrasound artifacts by clinicians can be inconsistent. Artificial intelligence (AI) may perform this task more consistently.

**Research Question:** Can AI characterize lung ultrasound artifacts similarly to humans, and can AI interpretation be corroborated by clinical data?

**Study Design and Methods:** Lung sonograms (n=665) from a convenience sample of 172 subjects were prospectively obtained using a pre-specified protocol and matched to clinical and radiographic data. Three investigators scored sonograms for A-lines and B-lines. AI was trained using 142 subjects and then tested on a separate dataset of 30 patients. Three radiologists scored similar anatomic regions of contemporary radiographs for interstitial and alveolar infiltrates to corroborate sonographic findings. The ratio of oxyhemoglobin saturation:fraction of inspired oxygen (S/F) was also used for comparison. The primary outcome was the intraclass correlation coefficient (ICC) between the median investigator scoring of artifacts and AI interpretation.

**Results:** In the test set, the correlation between the median investigator score and the AI score was moderate to good for A lines (ICC 0.73, 95% CI [0.53-0.89]), and moderate for B lines (ICC 0.66, 95% CI [0.55-0.75]). The degree of variability between the AI score and the median investigator score for each video was similar to the variability between each investigator’s score and the median score. The correlation among radiologists was moderate (ICC 0.59, 95% CI [0.52-0.82]) for interstitial infiltrates and poor for alveolar infiltrates (ICC 0.33, 95% CI [0.07-0.58]). There was a statistically significant correlation between AI scored B-lines and the degree of interstitial opacities for five of six lung zones. Neither AI nor human-scored artifacts were consistently associated with S/F.

**Interpretation:** Using a limited dataset, we showed that AI can interpret lung ultrasound A-lines and B-lines in a fashion that could be clinically useful.

## Introduction

Lung ultrasound (US) is used for real time identification and prognostication of lung pathology by clinicians at the bedside. Because of its ease of use and because it does not expose patients to ionizing radiation, this imaging modality has undergone explosive growth in intensive care units, emergency departments, and hospital wards to inform management decisions[1]. Lung ultrasound outperforms traditional radiographs in the diagnosis of some common lung pathologies such as cardiogenic pulmonary edema and pneumothorax[2,3].

Normally aerated lung attenuates the transmission of sound waves making it difficult to directly visualize disease pathology. Instead, the accurate interpretation of lung ultrasound relies on the characterization of reverberation artifacts that are generated at the interface of unaerated and aerated lung.

Lichstenstein designated over 40 lung ultrasound artifacts in his seminal work on thoracic sonography[4]. A-lines and B-lines are the artifacts most readily understood by practicing clinicians[1]. A-lines are generated by reflection of the ultrasound wavefront back and forth *between* the skin and the pleura. A-lines are present in healthy lung and some pathologic conditions, such as pneumothorax and emphysema. In contrast, B-lines are created by reverberations *within* the first millimeter of diseased but aerated lung. B-lines are most commonly seen when there is either interstitial edema or fibrosis. Increasing numbers of B-lines correspond to increasing disease severity[5,6].

The collage of reverberation artifacts in a lung ultrasound encodes important diagnostic information. However, the interpretation of this collage is subjective and only semi-quantitative leading to inconsistencies in interpretation, even among ultrasound fellowship-trained clinicians[7].

Because artificial intelligence systems (AI) can handle many more input variables, they have been shown to outperform humans in complex tasks such as the interpretation of mammograms[8]. We therefore hypothesized that an AI network could be trained to decode lung ultrasound artifacts in a fashion similarly to humans. To test this hypothesis we prospectively enrolled patients in a study investigating the correlations among human and AI scoring of lung US. We corroborated AI sonographic findings with selected clinical and radiographic variables. The specific reverberation artifacts chosen were A-lines and B-lines.

## Materials and Methods

This was a prospective, observational study conducted on a convenience sample of 172 adult patients admitted to a university-affiliated hospital from January 2021 to February 2022. The protocol was approved by the local institutional review board (LSU IRB#1509). All patients admitted to the study hospital over 18 years of age were eligible to participate. Written informed consent was obtained prior to the performance of any study related procedures.

### Ultrasound Protocol

Sonographers consisted of a Pulmonary/Critical Care attending, Pulmonary/Critical Care fellow, and five residents. All sonographers received specific training on the technique of obtaining quality lung ultrasounds. Training consisted of 2 hours of independent, directed learning using an accredited lung ultrasound educational product and 6 hours of didactic training involving lung ultrasound acquisition (S1 Appendix).

Patients were scanned with a point-of-care ultrasound system (X-Porte, Fujifilm Sonosite, Bothell, WA), using a linear array probe (HFL38xp/13-6 MHz) and the following presets: depth 6 cm, near field gain 0%, far field gain 100%, mechanical index 0.5, tissue index 0.2, tissue harmonics off. Patient’s were scanned in the sitting (preferred), or semirecumbent position at three points on each side: the 2nd intercostal space at the mid-clavicular line, 4th intercostal space at the mamillary line, and 5th intercostal space at the posterior axillary line (Figure 1) similar to the BLUE protocol (BLUE points)[9]. The probe was placed in the intercostal space and oriented parallel to the ribs. Six second clips were obtained at each point.

**Figure 1.**
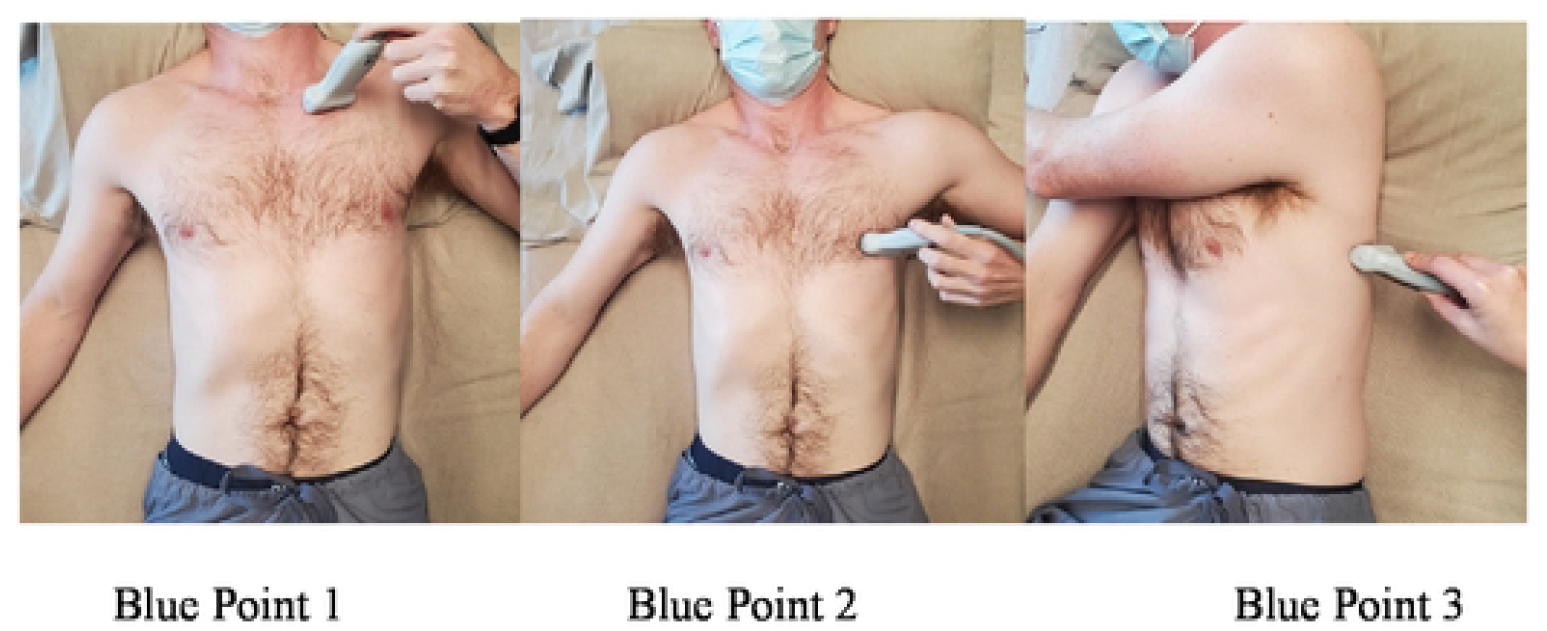
BLUE points of one hemithorax.

### Imaging Interpretation

Sonograms were scored by two physicians and one research staff member (BD, TF, GG), each blinded to clinical data. These investigators received 6 hours of explicit training on lung ultrasound interpretation using accredited training material (S1 Appendix)[10] and had at least an additional 2 years of research experience in this subject matter. Each sonogram was scored for the presence and character of A-lines and the quantity of B-lines on an ordinal scale (Table 1).

**Table 1.**
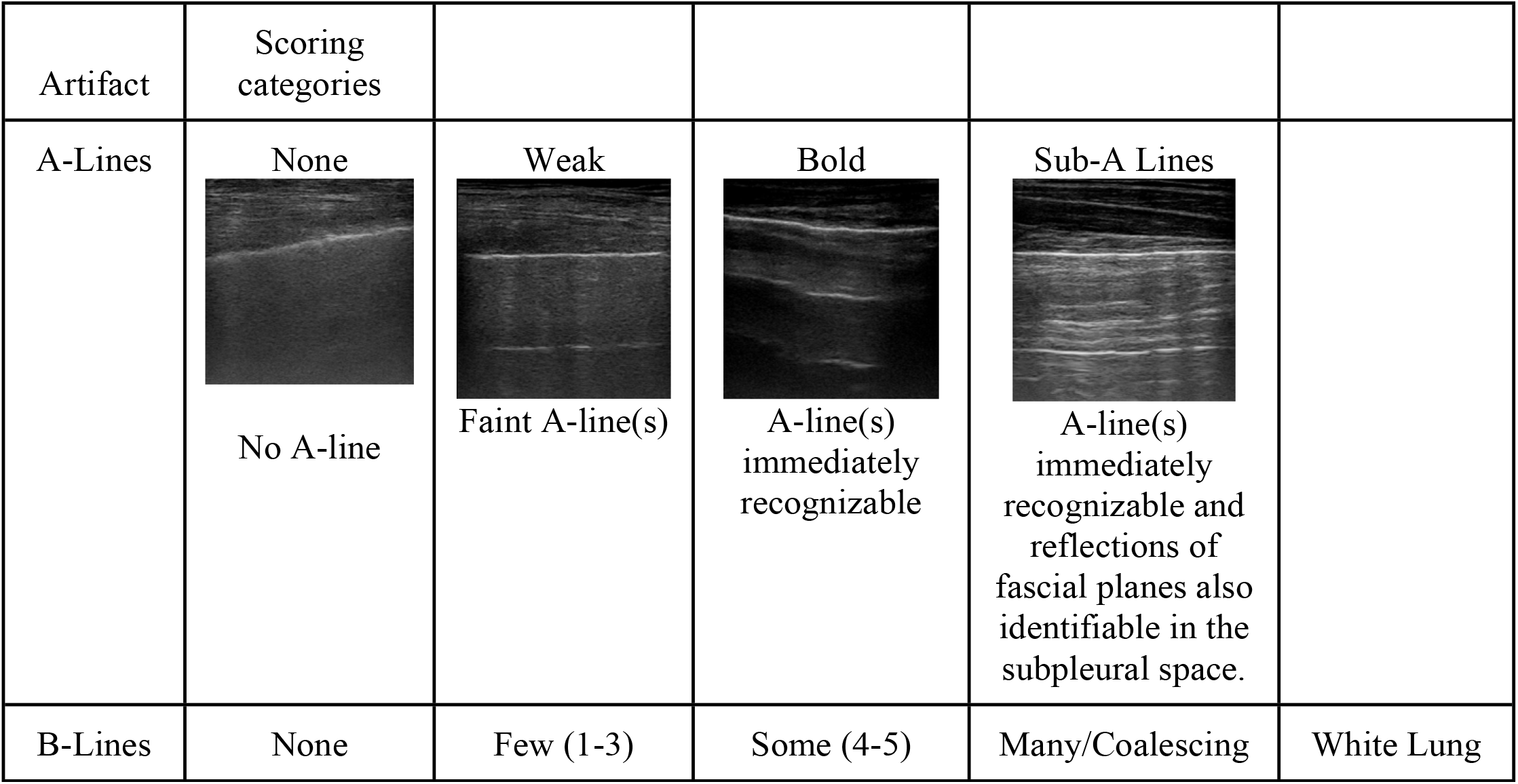
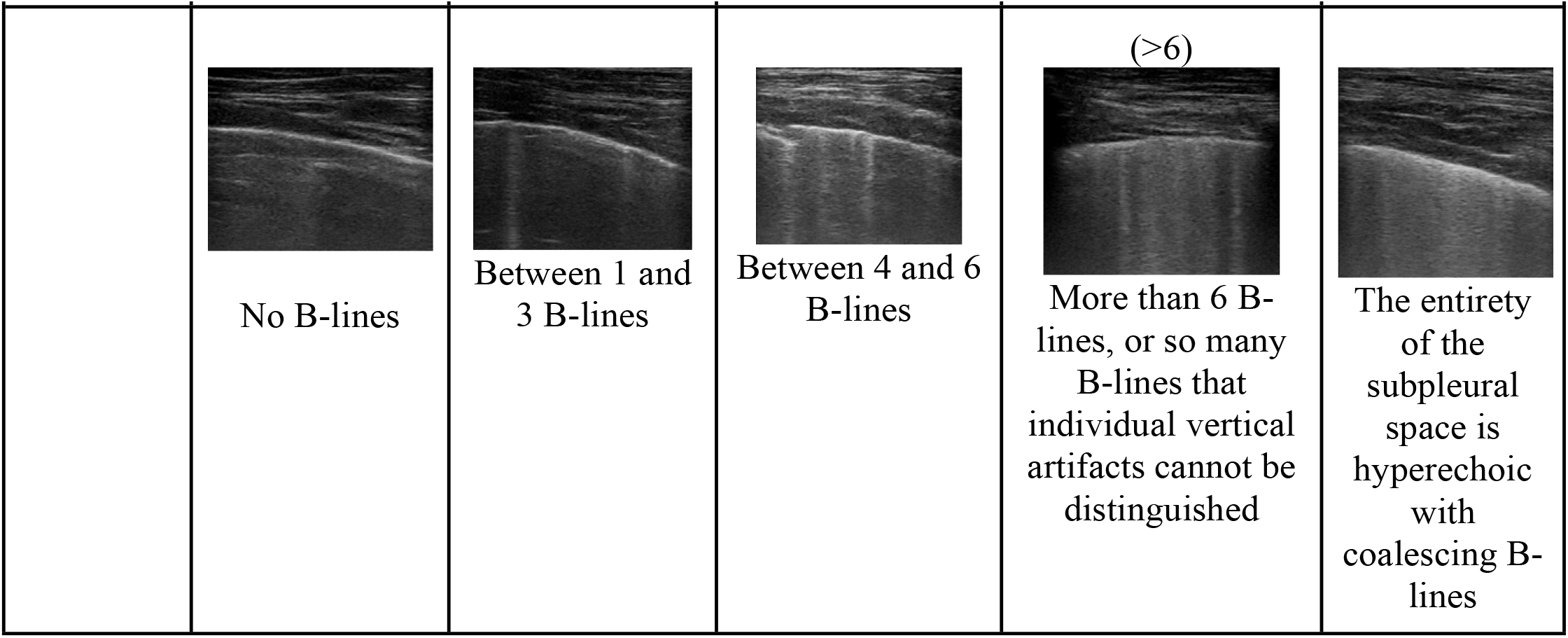
Lung Ultrasound Artifact Scoring

Three staff radiologists blinded to the clinical data independently scored the digital chest radiograph in the test set closest in time to the ultrasound exam. Radiographs obtained less than 24 hours from ultrasound acquisition were included. Radiographs were scored for the degree of interstitial and alveolar opacities at 6 different lung zones similar anatomically to the BLUE points (Figure 2)[11]. Each radiographic sextant was scored on a scale of 0 (no infiltrate) to 3 (dense infiltrates) for both interstitial and alveolar infiltrates using an electronic slider to provide a continuous variable[11]. To improve reproducibility, prior to scoring, each radiologist had obtained explicit instruction and had trained using the scoring system on a separate data set.

**Figure 2.**
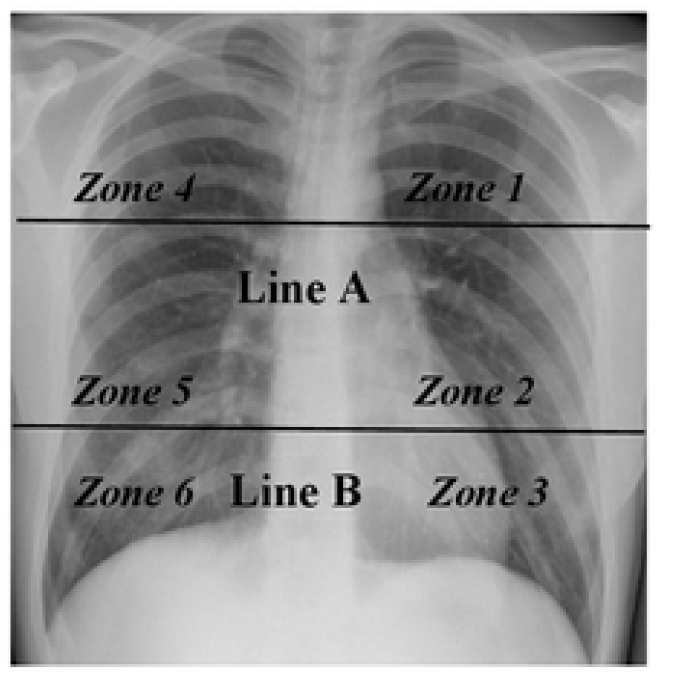
Chest radiograph scoring: Each chest radiograph was divided into 6 anatomical zones corresponding to the 6 ultrasound BLUE points. Two lines separate the thorax in the transverse plane, and the spinous process divides it sagittally to form six lung zones. Line A is drawn at the level of the inferior wall of the aortic arch. Line B is drawn at the level of the inferior wall of the right inferior pulmonary vein.

### Artificial Intelligence Network

A previously published artificial intelligence neural network which has been used to analyze lung ultrasound artifacts[12,13] by employing a Temporal Shift Module (TSM)[14] was trained using 485 ultrasound clips from 142 research subjects. The previously published model characterized A and B line artifacts with an accuracy of 76.4% and a precision of 70.8%[13]. The TSM model is video-based model that jointly analyzes a group of frames belonging to a video clip in order to simultaneously predict A-lines and B-lines[14]. Such a video-model is better suited to detect transitory features (B-lines) rather than frame-based models[15,16] that only use a single frame for analysis.

In the present study, no crossover existed between patients in the training set and those in the test set. Subjects in the training and test sets had sonograms conducted in a similar fashion to that outlined as above. For AI training, each sonogram clip was pre-labeled by one of the investigators (BD, TF, or GG) using an annotator that captured the predominant artifacts. The training involved exposing AI to the labeled video clip, as has been described previously[14].

Once trained, AI was tasked with interpreting a separate test set of 180 unlabelled clips from 30 patients for A-lines and B-lines. For each clip, AI predicted a probability that selected A-line patterns and selected B-line patterns would be present. Any A-line or B-line pattern with a probability greater than 50% was scored as being present. For example, in a single clip, AI may produce a probability that weak A-lines were present (40%), bold A-lines were present (60%), etc. If two or more A-line patterns were scored as being present, the bolder descriptor for the pattern was chosen. For example, if weak and bold A-lines were scored as being present (had probability scores greater than 50%), the bold A-line option was chosen. In another example, if “few” and “many/coalescing” B-lines were scored as being present, the clip was scored as having “many/coalescent” B-lines. This determination was made *a priori*, and is consistent with clinician scoring. It was not anticipated prior to data interpretation that there would be clips where AI was unable to identify a B-line pattern with a probability greater than 50%. In these instances we chose the B-line pattern with the highest probability, even if that probability was less than 50%.

### Clinical Data

In the test set, the following demographic and clinical descriptors were obtained at the time of each exam: age, gender, admission location, arterial oxygen saturation/fraction of inspired oxygen (S/F), BMI, final diagnosis at discharge, and NIH ordinal scale[17]. S/F was determined using pulse oximetry performed concurrently with the lung US. Diagnosis and NIH ordinal scale was established by reviewing the primary team’s documentation, contributing lab and imaging studies, and response to treatments. Method of diagnosis for specific conditions is included in the online supplement(S1 Table).

### Statistical Analysis

The primary outcome was the intraclass correlation coefficient (ICC) between the median investigator artifact interpretation and the AI artifact interpretation. Secondary outcomes included inter-rater reliability of lung artifact interpretation among investigators, and interrater reliability among chest radiograph scoring among reading radiologists, both determined by the intraclass correlation coefficients. All ICCs were calculated using a two-way effects, absolute agreement, and single rater model and reported with 95% confidence intervals.

External validation was achieved through comparison of AI and investigator artifact interpretation with radiographic characteristics of pulmonary disease, as well as oxygenation as measured by S/F ratio. These relationships were quantified with an analysis of variation (ANOVA). Data were reported as p-values and effect quantification via η^2^ with statistical significance defined as a p-value less than 0.05. Prior to conducting the ANOVA, Levene’s test of equality was used to confirm homoscedasticity. Relationships between the ultrasound artifact interpretation and the clinical data were visualized using box plots. All analyses were conducted using R version 4.1.3 (R Core Team for Statistical Computing, Austria).

## Results

Clinical data for research subjects in the test set are shown in Table 2. The average patient age was 66 years; the majority were admitted to an ICU (40%); and there was a relatively equal mix of men (53%) and women (47%). The most common diagnoses were decompensated heart failure (n=7) and COVID-19 pneumonia (n=7), with bacterial pneumonia, chronic obstructive lung disease (COPD) exacerbations, pleural effusion, and interstitial lung disease making up a minority of diagnoses (Table 3).

**Table 2.** Demographic and Clinical Information *a*:Median (25%, 75%); n (%)

| **Characteristic** | N=30<sup>a</sup> |
| --- | --- |
| **Age (years)** | 66 (51,78) |
| **Gender** |  |
| Female | 14 (47%) |
| Male | 16 (53%) |
| **Disposition** |  |
| Inpatient Ward | 12 (40%) |
| Intensive Care Unit | 18 (60%) |
| **BMI** | 28 (26, 32) |
| **S/F** | 330 (240, 372) |
| **NIH Ordinal Scale** |  |
| 3 | 2 (6.7%) |
| 4 | 2 (6.7%) |
| 5 | 17 (57%) |
| 6 | 7 (23%) |
| 7 | 2 (6.7%) |

*a*:Median (25%, 75%); n (%)

**Table 3:** Frequency table for diagnosis for enrolled patients in the test set.

| Diagnosis* | Number of Patients |
| --- | --- |
| Heart Failure Exacerbation | 7 |
| COVID-19 Pneumonia | 7 |
| Pleural Effusion | 5 |
| Interstitial Lung Disease | 5 |
| COPD Exacerbation | 4 |
| Bacterial Pneumonia | 2 |
| Other | 2 |

\* Multiple diagnoses could exist in the same patient

### Inter-rater agreements

Among clinicians there was moderate to good agreement overall in A-line pattern description (ICC= 0.75 [95% CI: 0.64-0.83]), and moderate agreement in B-line pattern description (ICC= 0.71 [95% CI 0.58-0.79]). AI scoring of A-lines had moderate to good agreement with the median human A-line score using intraclass correlation coefficient (ICC= 0.73 [95% CI 0.53-0.84]). AI scoring of B-lines also showed moderate agreement with median human scoring (ICC= 0.66 [95% CI 0.55-0.75)](Tables 4, 5).

**Table 4.** ICC among human and AI scoring of A-lines [95% CI]

| **Anatomic Location** | **ICC among Human Scoring** | **ICC between AI and Median Human Scoring** |
| --- | --- | --- |
| **BLUE 1 (n=30)** | **0.92** [0.85-0.96] | **0.88** [0.76-0.94] |
| **BLUE 2 (n=30)** | **0.70** [0.53-0.83] | **0.82** [0.66-0.91] |
| **BLUE 3 (n=30)** | **0.63** [0.36-0.80] | **0.62** [0.1-0.85] |
| **BLUE 4 (n=30)** | **0.74** [0.58-0.85] | **0.82** [0.63-0.91] |
| **BLUE 5 (n=30)** | **0.80** [0.63-0.90] | **0.68** [0.43-0.83] |
| **BLUE 6 (n=30)** | **0.52** [0.24-0.73] | **0.43** [0.02-0.70] |
| **Mean (n=180)** | **0.75** [0.64-0.83] | **0.73** [0.53-0.84] |

**Table 5.**
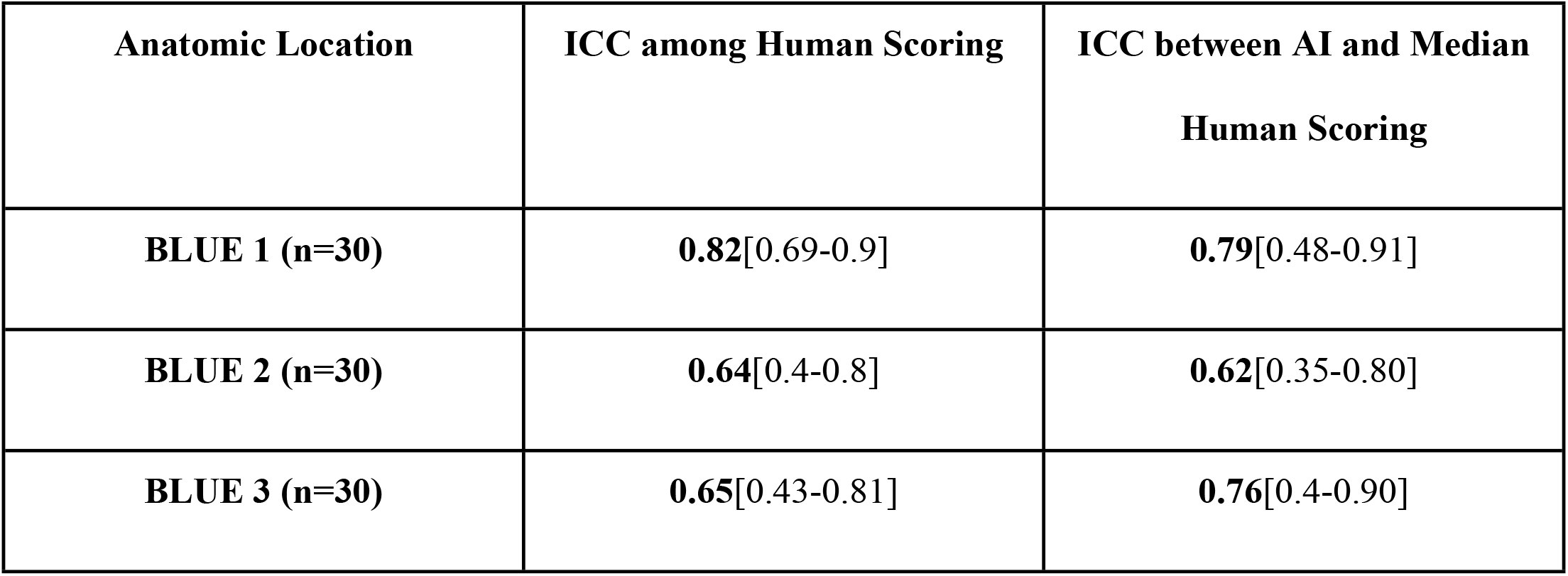
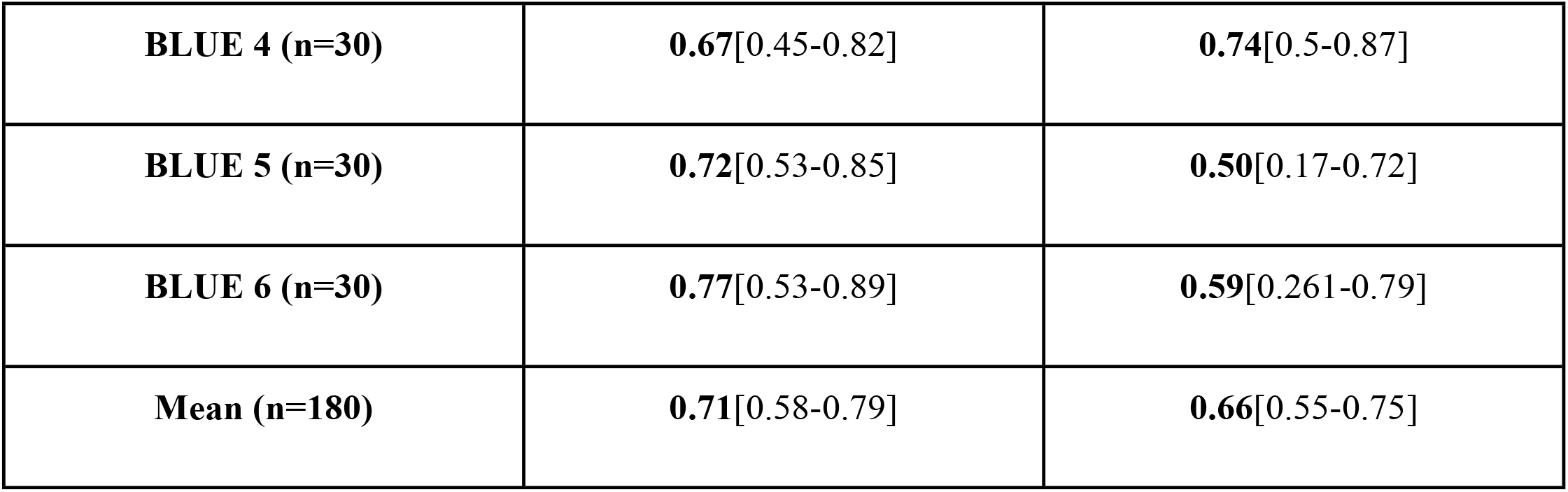
ICC among AI and median human US scoring of B-lines [95% CI]

| **Anatomic Location** | **ICC among Human Scoring** | **ICC between AI and Median Human Scoring** |
| --- | --- | --- |
| **BLUE 1 (n=30)** | **0.82**[0.69-0.9] | **0.79**[0.48-0.91] |
| **BLUE 2 (n=30)** | **0.64**[0.4-0.8] | **0.62**[0.35-0.80] |
| **BLUE 3 (n=30)** | **0.65**[0.43-0.81] | **0.76**[0.4-0.90] |
| **BLUE 4 (n=30)** | **0.67**[0.45-0.82] | **0.74**[0.5-0.87] |
| **BLUE 5 (n=30)** | **0.72**[0.53-0.85] | **0.50**[0.17-0.72] |
| **BLUE 6 (n=30)** | **0.77**[0.53-0.89] | **0.59**[0.261-0.79] |
| **Mean (n=180)** | **0.71**[0.58-0.79] | **0.66**[0.55-0.75] |

To more directly compare the variability between AI and each investigator, AI was considered as a separate investigator. Then for each artifact pattern, the variability between AI and the median score of all investigators was compared to the variability of each investigator and the overall median. For A-lines, the ICC between each investigator and the median score for each clip ranged from 0.74 (TF vs median) to 0.83 (BD vs median). AI had similar variability with an ICC of 0.74 when compared to the median investigator score. For B-lines, investigator variability ranged from 0.6 (ICC between TF vs median of AI and the other investigators) to 0.75 (ICC between GG vs median of AI and the other investigators) while AI variability was 0.65 (ICC between AI vs median investigator score) (table 6).Thus AI performed within the range of human scoring for the detection of specific A-line and B-line artifact patterns.

**Table 6.** Variance shown as ICC [95% CI] between Human and AI scoring of Artifacts versus Median

|  | **TF vs median** | **BD vs median** | **GG vs median** | **AI vs median** |
| --- | --- | --- | --- | --- |
| **A-Lines** | **0.74** [0.48-0.85] | **0.83** [0.76-0.88] | **0.77** [0.6-0.85] | **0.74** [0.53-0.84] |
| **B-Lines** | **0.6** [0.26-0.77] | **0.71** [0.61-0.78] | **0.75** [0.64-0.83] | **0.65** [0.54-0.73] |

Furthermore, there was not a significant difference in interrater reliability in any one disease state over another, although AI scoring was most similar to humans in scoring sonograms of patients with COVID and least similar in patients with COPD (S2 Table). Although many clinicians use radiology to inform patient care of respiratory diseases, the ICC among radiologist scoring of both interstitial and alveolar infiltrates was moderate to poor (Table 7).

**Table 7.** Inter-rater reliability among radiologists for interstitial and alveolar opacities shown as ICC [95% CI]

| Anatomic Location | Interstitial Scoring | Alveolar Scoring |
| --- | --- | --- |
| **BLUE 1 (n=30)** | **0.30** [0.09-0.53] | **0.24** [0.04- 0.47] |
| **BLUE 2 (n=30)** | **0.65** [0.47-0.79 ] | **0.40** [0.18-0.61] |
| **BLUE 3 (n=30)** | **0.56** [0.35 -0.73] | **0.24** [0.04-0.47] |
| **BLUE 4 (n=30)** | **0.60** [0.4-0.76] | **0.00** [-0.16-0.23] |
| **BLUE 5 (n=30)** | **0.69** [0.52-0.82] | **0.35** [0.13-0.57] |
| **BLUE 6 (n=30)** | **0.44** [0.22-0.65] | **0.36** [0.10-0.60 ] |
| **Mean (n=180)** | **0.59** [0.52-0.82] | **0.329** [0.07-0.58] |

### Comparisons with Clinical Data

A statistically significant association (p<0.05) was found between the density of interstitial opacities in corresponding chest radiographs and number of B-lines counted by AI in five of six anatomic lung zones using an ANOVA with a large effect size (η^2^ range: 0.21-0.47). Similarly, the density of interstitial opacities was associated with investigator-scored B-lines (S4,5 Tables). No statistically significant association was found between the density of interstitial opacities and the strength of A-lines as scored by either AI or investigators using an ANOVA (S6,7 Tables). Box plots of B-lines versus interstitial opacities demonstrated an increased B-line number by both human and AI scoring in lung zones with denser interstitial markings on chest radiographs (Figure 3).

**Figure 3.**
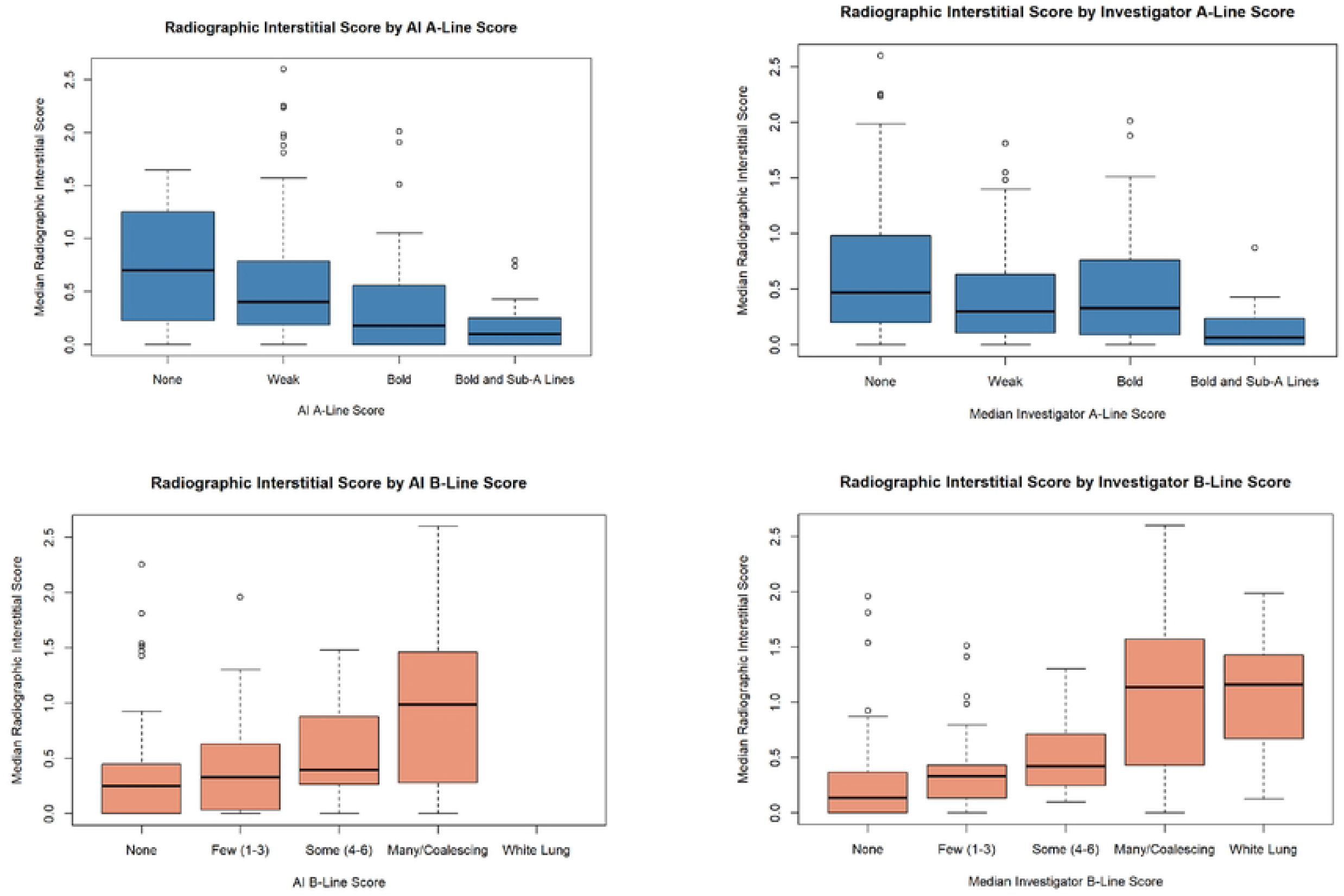
Box plots of radiographic interstitial score and artifact interpretation are shown. Median interstitial score determined by radiologists are shown on the Y-axis. Videos scored for the character of the A-line or number of B-lines by either AI or the median human score. The black bar indicated the median radiographic interstitial scoring. The colored bar represents the 25th and 75th percentile. The extent of the whiskers indicate the 95% confidence interval.

Three of six lung zones had a statistically significant association (p<0.05) between the degree of alveolar opacities and AI-scored A-lines, with a large effect size (η^2^ range: 0.25-0.29). In comparison, two of six lung zones showed a statistically significant association between the degree of alveolar opacities on chest radiographs and investigator-scored A-lines (S9,10 Tables). No statistically significant association was found between the density of alveolar opacities and the number of B-lines on sonograms scored by either AI or investigators (S11,12 Tables).

A statistically significant association was found between oxygenation via S/F and AI-scored A-lines in three of six lung zones using an ANOVA (S13,14 Tables), and also demonstrated visually by box plots (Figure 4). There was no statistically significant association between S/F and the brightness of A-lines as scored by humans (S15,16 Tables).

**Figure 4.**
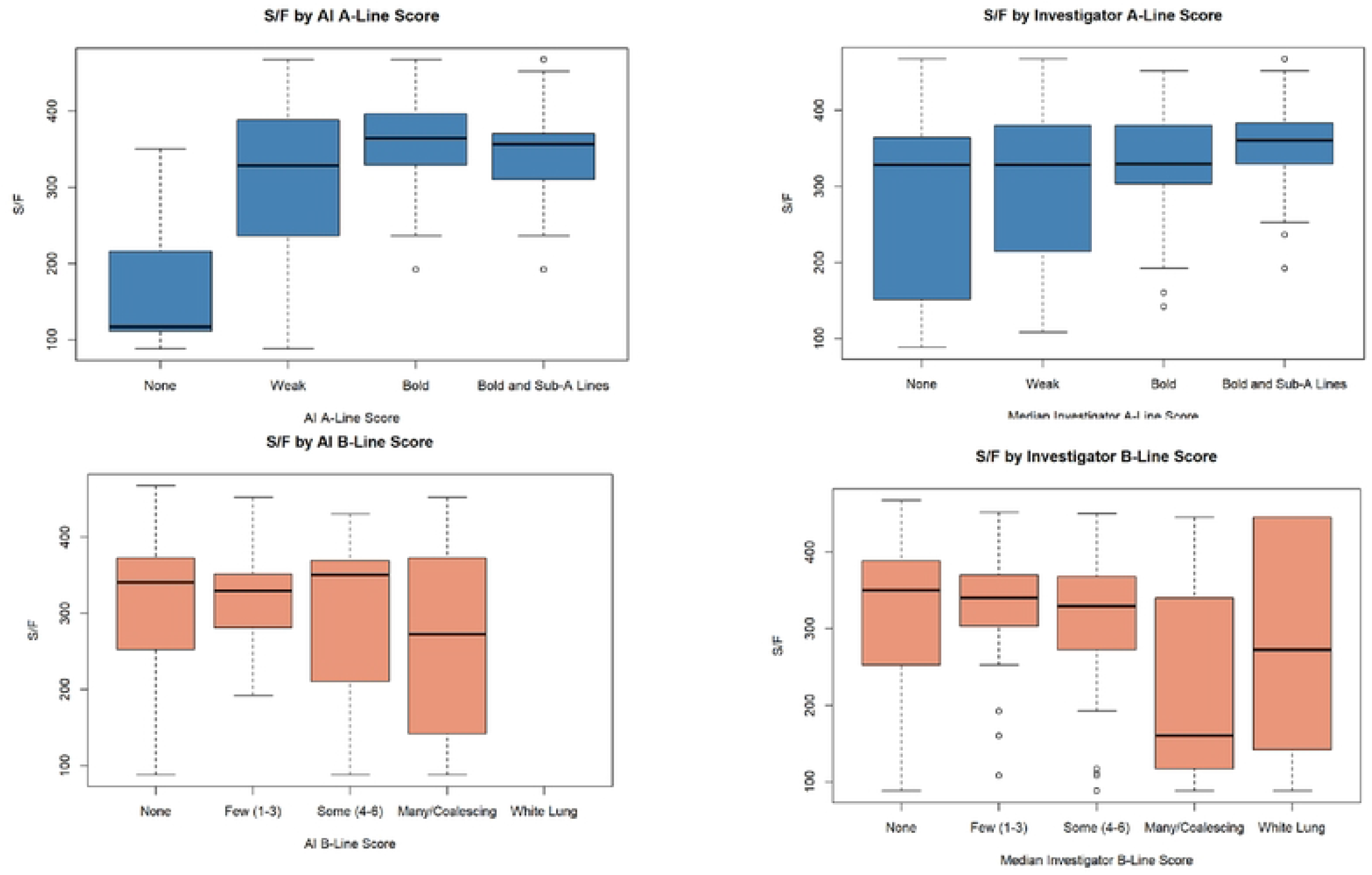
Box plots of lung function quantified by S/F and artifact interpretation are shown. S/F for each research subject is shown on the Y axis. Human and AI scoring of A-lines and B-lines are shown.The black bar indicated the median S/F for all videos scored as having each character of A-line or number of B-lines. The colored bars represent the 25th and 75th percentiles. The whiskers indicate the 95% confidence interval.

## Discussion

Human sonographers show significant variability in the scoring of lung ultrasound artifacts. In spite of this unwanted scoring heterogeneity, point-of-care ultrasound is commonly used to inform patient care decisions. We also observed variability in our human scoring of lung ultrasound artifacts, furthermore the degree of variability was in line with existing evidence on this topic[7,18].

Unlike human scoring, a fully trained AI network holds the promise of yielding highly reproducible results. In the present study, we observed a moderate correlation between AI and investigator interpretation of A-lines, indicating that AI interpreted clips similarly to investigators for this artifact. There was a weaker correlation between AI and investigator scoring of B-lines, although the degree of correlation was in line with existing evidence on this topic[16,19]. In composite, these data suggest that AI trained on a relatively small data set can interpret A-line and B-line artifacts within the range of human interpretation.

Chest radiographs are a commonly ordered imaging modality in hospitalized patients. It is clear, however, that like other imaging modalities, there is significant variability in how chest radiographs are interpreted[20]. When compared to the degree of variability among radiologists interpreting interstitial and alveolar infiltrates, human sonographers and AI scored A-line and B-line artifacts with lower interrater variability.

Previous studies have shown a positive correlation between increasing interstitial infiltrates and the number of B-lines[5,6]. We observed a similar relationship between the density of interstitial infiltrates and the number of B-lines scored by both humans and AI. It is notable that AI did not score any clips in the test set as having the highest B-line severity (3 or “white lung”), perhaps because too few clips of this severity were included in the training set. However, ultrasound clips scored by clinicians as having the highest severity B-line score had wide confidence intervals, which may indicate that this ultrasound finding is not a reliable indicator of worsening interstitial disease.

It was less clear how alveolar opacities on chest radiographs would correlate with lung ultrasound interpretation. In this dataset, the degree of alveolar opacification, as adjudicated by radiologists, was inversely correlated with the boldness of A-lines as interpreted by AI in three of six lung zones. Somewhat surprisingly, the B-line artifact was not a reliable predictor of alveolar infiltrates on chest radiographs (S12,13 Tables).

Artificial intelligence neural networks have previously shown the ability to differentiate normal from abnormal lung sonograms, identifying, for example, an A-line predominant versus B-line predominant clip[21]. AI has also been shown to improve novice lung sonographers interpretation[19]. AI systems have previously been able to characterize multiple lung ultrasound artifacts simultaneously compared to a human standard[22]. These studies often do not attempt to analyze AI artifact identification beyond its similarity to human interpretation[23].

There are two novel aspects to the present study that extend previous observations. First, we tasked AI with characterizing more artifacts in more detail than previous studies. Second, we matched AI ultrasound artifact interpretations not only to human interpretation of the same sonogram, but also to that of an entirely different radiographic modality, as well as to physiologic data. We demonstrated that AI scoring compared favorably to human interpretation of sonograms, and that it correlated with other radiographic data. This provides added clinical relevance to AI interpretation of lung ultrasound artifacts.

Several protocols have been used to obtain thoracic sonograms for research studies, with variations in probe selection, orientation, and depth settings[24,25]. Of available probes, a high frequency, linear array probe was chosen for the present study because it allows enhanced resolution of the pleural line and subpleural structures[26]. The probe was placed in the intercostal space and oriented parallel to the ribs to allow visualization of a larger area of pleural surface uninterrupted by rib shadow[26,27]. This orientation also allowed for continuous contact with the skin along the entire length of the probe. Six cm was the maximum depth setting available for the probe used in this study.

### Limitations

Our study has limitations. First, it was conducted on a small convenience sample of patients from a single center, and may not be applicable to broader patient populations with more diverse pathologies. Second, all patients were scanned using an explicit protocol involving a single ultrasound model and probe, which may limit its generalizability to other ultrasound manufacturers, probes, and scanning techniques. Third, there was a limited number of sonographers and human interpreters, and the experience beyond the explicit training received as part of this experiment is not uniform. Fourth, the training set was over-represented with clips taken at the first and fourth BLUE points as opposed to clips more caudally located on the thorax, which may have contributed to the lower reliability in AI rating at these anatomic locations. Fifth, only two artifacts were measured in this study, and some patients’ pathology cannot be characterized using these artifacts alone, such as those with pleural effusions. Sixth, we recognize that in very obese patients, there may be more three centimeters of soft tissue between the skin and the pleural line which might have limited our ability to detect A-line artifacts. In this uncommon occurrence, we attempted to compress the probe against the skin until the skin to pleural distance spanned less than 3 cm. Seventh, imaging studies were interpreted by only six investigators (three for ultrasound, three for radiographs), which limits the statistical validity of variability among humans. Despite these limitations, this study represents encouraging evidence of the potential for machine learning to accurately characterize ultrasound artifacts with clinical implications.

## Conclusions

In this prospective, observational study of a small convenience sample of adults admitted to a university affiliated hospital, we demonstrate that an artificial intelligence network can be trained to identify and characterize A-line and B-line artifacts within the range of variability of human interpreters. We corroborate these interpretations with radiographic and clinical comparators that show AI interpretation of B-lines is associated with degree of interstitial disease.

## Data Availability

Data cannot be shared publicly because of stipulations of the IRB approved protocol. Data are available from the LSU IRB for researchers who meet the criteria for access to confidential data.

## Abbreviations

AI: Artificial intelligence neural network
BLUE 1: Point 1 on left side of thorax
BLUE 2: Point 2 on left side of thorax
BLUE 3: Point 3 on left side of thorax
BLUE 4: Point 1 on right side of thorax
BLUE 5: Point 2 on right side of thorax
BLUE 6: Point 3 on right side of thorax
COPD: Chronic Obstructive Pulmonary Disease
ICC: Intraclass correlation coefficient
US: Ultrasound
S/F: Oxyhemoglobin saturation divided by fraction of inspired oxygen

## Acknowledgements

BD had full access to all of the data in the study and takes responsibility for the integrity of the data and the accuracy of the data analysis. TF, LH, VP, GG, DS, FB, RD, HT, PL, DM, KZ, and BD acquired data. TF, GG, AK, JG, and BD contributed to the conception and design of the study. TF and GG performed statistical analyses. All authors participated in the interpretation of the data, provided critical feedback and final approval for submission, and took responsibility for the accuracy, completeness, and protocol adherence of data and analyses.

